# Diagnostic Accuracy of FDA Authorized Serology Tests to Detect SARS-CoV-2 Antibodies: A Systematic Review and Meta-analysis

**DOI:** 10.1101/2020.10.07.20208553

**Authors:** Rajesh Pandey, Anand Gourishankar

**Affiliations:** Division of Neonatology, McGovern Medical School, University of Texas Health Science Center at Houston, Houston, TX, USA; Division of Pediatric Hospital Medicine, Children’s National Hospital, The George Washington School of Medicine and Health Sciences, Washington DC, USA

## Abstract

**Importance:** Serology tests are diagnostic and complementary to molecular tests during the COVID-19 pandemic.

**Objective:** To evaluate the diagnostic accuracy of FDA authorized serology tests for the detection of SARS-CoV-2 infection.

**Data sources:** A search of MEDLINE, SCOPUS, CINAHL Plus, and EMBASE up to April 4, 2020, was performed to identify studies using the “COVID 19 testing” and “meta-analysis.” FDA website was accessed for the list of tests for emergency use authorization (EUA).

**Study Selection:** Manufacturer reported serology tests published in the FDA website were selected. Two reviewers independently assessed the eligibility of the selected reports.

**Data extraction and synthesis:** The meta-analysis was performed in accordance with the PRISMA guidelines. A bivariate analysis using the “random-effects model” was applied for pooled summary estimates of sensitivity, specificity, and the summary receiver operating characteristic curves.

**Main outcomes and measures:** The primary outcome was the diagnostic accuracy of the serology test for detecting SARS-CoV-2 infection. Subgroup analysis of the diagnostic accuracy with lag time between symptom onset and testing were studied.

**Results:** Seven manufacturer listed reports were included. The pooled sensitivity was 87% (95% CI, 78% - 93%), the pooled specificity was 100% (95% CI, 97% - 100%), and the area under the hierarchical summary receiver operating characteristic curve was 0.97. At ≤ 7 days, sensitivity was 44% (95% CI, 21% - 70%), and for 8-14 days, sensitivity was 84% (95% CI, 67 % - 94%).For blood draws ≥ 15 days after the onset of symptoms, sensitivity was 96% (95% CI, 93% - 98%). Heterogeneity was substantial, and the risk of bias was low in this analysis.

**Conclusions and relevance:** FDA authorized serology tests demonstrate high diagnostic accuracy for SARS-CoV-2 infection (certainty of evidence: moderate). There is a wide variation in the test accuracy based on the duration between the onset of symptoms and the tests (certainty of evidence: low).

**Key– points:** 

**Questions:** What is the pooled diagnostic accuracy of FDA authorized serology tests to detect SARS-CoV-2 antibodies?

**Findings:** In this systematic review and meta-analysis of seven reports from FDA authorized serology tests to detect antibodies against SARS-CoV2 antibodies (3336 patients/ samples) pooled sensitivity was 87%, and pooled specificity was almost 100%. There was a wide variation in test performance based on the duration between the onset of symptoms and the tests.

**Meaning:** FDA authorized tests are highly accurate to detect antibodies against SARS-CoV-2 antibodies if tests are performed under a similar condition, as presented in the original report. There is a wide variation in the test performance based on the time interval between the onset of symptoms to the tests.

## Background

Coronavirus disease (COVID-19) is a current global threat, and diagnostic tests are critical. COVID-19 was discovered in Hubei Province, China, in December 2019, caused by a pathogen of unknown origin,^1,2^ which was subsequently named severe acute respiratory syndrome coronavirus 2 (SARS-CoV-2). COVID-19 has currently caused a pandemic. ^3, 4^ The genome sequence of SARS-CoV-2 was added to the GenBank sequence repository on January 10, 2020, ^2,5^ which enabled laboratories across the world to develop and use reverse transcription-polymerase chain reaction (RT-PCR) test for the diagnosis. Both the World Health Organization (WHO) and the Centers for Disease Control and Prevention (CDC) recommend an RT-PCR for the diagnosis of SARS-CoV-2 infection. ^6,7^

SARS-CoV-2 is a new pathogen in humans, so there are no antibodies found in a person not exposed to this virus. ^7^ The rise in antibody titer between acute and convalescent-phase sera tested in parallel is definite evidence of the viral infection. ^7^ Seroconversion after SARS-CoV-2 infection appears as early as the fourth day after the onset of symptoms, ^8^ but can be delayed beyond 15 days. ^9^ Previous publications have supported the use of serological tests for the detection of SARS-CoV-2 infection. ^9,10^ WHO has affirmed the use of seroconversion or an increase in antibody titer for the diagnosis of SARS-CoV-2. ^7^ However, the CDC does not recommend serological tests for diagnosis of SARS-CoV-2. Nonetheless, the Food and Drug Administration (FDA) has authorized emergency use authorizations (EUA) for diagnostic serological tests for COVID-19 ^6, 11^ as a valuable tool for the management of this pandemic.

Serological tests offer several advantages including (1) contact tracing; (2) serologic surveillance; (3) identification of those who have already had the virus infection and thus may (if there is protective immunity) be immune; ^12^ (4) to identify individuals as a potential source for convalescent serum treatments (currently an experimental treatment or prophylaxis); ^13, 14^ (5) to do epidemiological researches; (6) theoretically to diagnose a person in a late stage of disease when RT-PCR may be false negative; ^15^ and (7) evaluation and development of a vaccine. ^15,16^

With the novel Coronavirus surge, testing brings challenges with variability in performance, including sensitivity and specificity. ^15^ Therefore, the purpose of this review was to evaluate the diagnostic accuracy of the FDA authorized diagnostic serological tests for SARS-CoV-2 infection.

## Methods

### Literature search

We used the FDA website to search for all FDA authorized, under emergency use authorization (EUA), in-vitro serology diagnostic tests for SARS-CoV-2. We performed a search for systematic reviews up to April 4, 2020, using the term “COVID 19 testing” and “meta-analysis” in the following databases: MEDLINE, CINAHL Plus, EMBASE, and SCOPUS. This search was limited to publication in the “English” language.

### Inclusion criteria

We included all FDA authorized diagnostic serological tests (before April 26, 2020) available to use in the United States of America. We collected information from the authorization reports, either the manufactures’ instructions/package insert. ^11^ Inclusions also had the following criteria: index test must include qualitative detection of anti-SARS-CoV-2 antibodies [Immunoglobulin G (IgG) or Immunoglobulin M (IgM), or both], and target condition must be confirmed by RT-PCR (the reference standard).

### Exclusion criteria

(1) Serology tests not authorized by the FDA; (2) serology tests not authorized for diagnostic use; (3) molecular tests authorized by the FDA.

### Data extraction and quality assessment

Two authors (RP and AG) reviewed all eligible reports and extracted the data independently. Disagreements between the researchers were resolved by consensus. We extracted the following data: (1) manufacturer of the test; (2) settings including sample size, disease status at the time of specimen collection, other clinical contexts available in the report; (3) information regarding test-assay methods, specimens used; and (4) data on a true positive, false positive, false negative and true negative. There was a complete agreement on the final data used in the analysis.

We assessed the methodological quality of each study by following the guidelines from the Cochrane Screening and Diagnostic Test Methods Group, a tool adapted from the QUADAS-2. The four domains assessed for risk of bias were patient selection, index test, reference standard, and flow and timing. We assessed the applicability concerns in the first three domains. In each domain, we answered the signaling questions with ‘Yes,’ ‘No,’ or ‘Unclear.’ For each domain, the risk of bias was judged as ‘Low,’ ‘High,’ or ‘Unclear’ risk. One review author assessed study quality, which a second review-author verified. We summarized the overall quality of evidence using the GRADE methodology recommended for diagnostic tests. The assessment of publication bias was not applicable based on our research design.

### Data synthesis and analysis

We constructed 2x 2 tables for all reports based on dichotomous data from the reference standard (positive or negative). We performed data analysis using methods described in the Cochrane Handbook of Diagnostic Test Accuracy (DTA) Reviews. ^17^ We created forest plots with 95% confidence intervals (CI) for sensitivity and specificity for each study using Review Manager 5. ^18^ Summary statistics of four sets of primary data, namely true positive (TP), false positive (FP), false negative (FN), and true negative (TN), were analyzed. Seroconversion, which determines the accuracy of tests, after SARS-CoV-2 infection depends on the time interval between the onset of disease and testing. ^9, 19, 20^ To address the test accuracy and variation in sensitivity of tests with the lag time, we performed subgroup analysis for tests done at (1) at or before 7days; (2) between 8-14 days and (3) at or after 15 days after the onset of symptoms. A bivariate random-effects model was used for summary effect size, and Area under curve (AUC) using R software version 3.2. ^21, 22^ This bivariate model takes account into binomial distribution for directly modeling sensitivity and specificity for within-study variations, and assumes bivariate normal distribution for between-study variation. ^23^ We used inverse variance when calculating the weights of individual studies. The DerSimonian-Laird estimator was used when calculating between-study variations for statistical assumptions and conservative estimates. The confidence interval was estimated by the Clopper-Pearson method. Heterogeneity was categorized as follows: not important (0% - 40%); moderate (30% - 60%), substantial (50% - 90%); and considerable (75% - 100%). ^24^ In addition to ROC, the Spearman’s correlation coefficient (Rho) between the sensitivity and false-positive rates was calculated for the presence of “cut-off” point (≥ 0.6). ^25^ GRADEpro software was used to create a summary of findings table. ^26^ Publication bias deferred in this analysis as we used manufacturer quoted reports published by the FDA.

## Results

The systematic literature search yielded no meta-analysis on serological tests for SARS-CoV-2 infection as of April 4, 2020. A total of 48 EUA summaries were identified from the FDA website accessed on April 25, 2020, of which seven reports which met eligibility criteria were included (eFigure 1 in the supplement).

### Characteristics of the included reports

All seven studies are exclusively obtained from the FDA website. Information was collected from the information for use (IFU) documents. The characteristics of the included reports are summarized in Table 1.

**Table 1:**
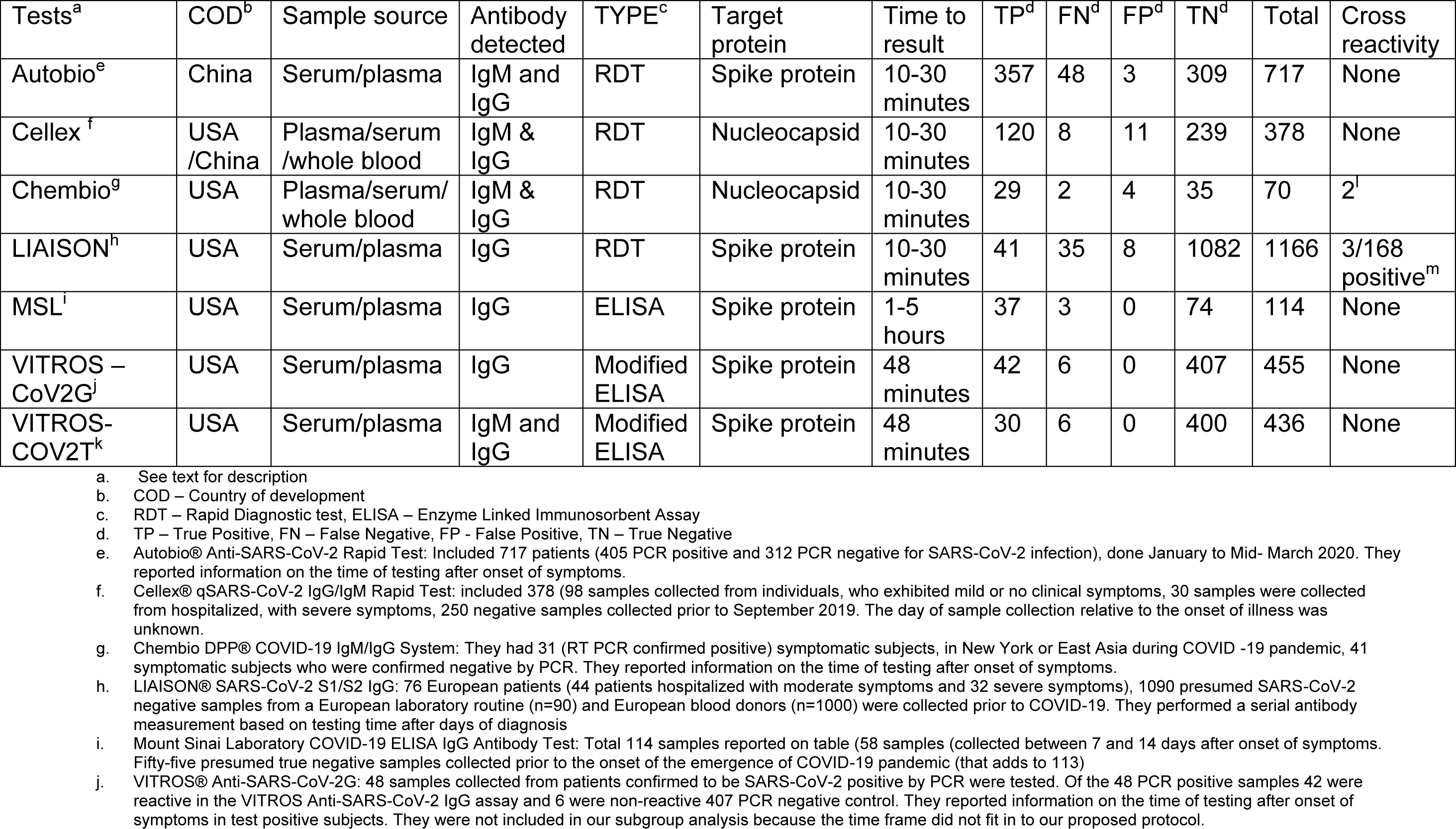
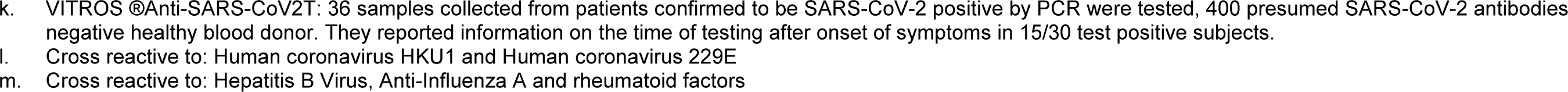
Characteristics of the reports included in the systematic review.

The sample sizes in each report ranged from 70 to 1166 (total n = 3336). Five reports used a qualitative detection of IgM, IgG, or both antibodies against the SARS-CoV-2, while the remaining two used qualitative detection for the detection of IgG only. Demographics such as age, ethnicity, gender were not available. Comorbidities of the patients who tested positive for SARS-CoV-2 infection were unknown. As the cases are rising rapidly, there was no information available regarding the prevalence of the disease.

There was an incomplete and variable reporting of the time interval between the onset of symptoms and the test. Autobio^®^ Anti-SARS-CoV-2 Rapid Test had information on the timing of the test relative to the onset of symptoms while Cellex^®^ qSARS-CoV-2 IgG/IgM Rapid Test did not have such information. Chembio DPP^®^ COVID-19 IgM/IgG System test had information on days of blood samples collected from symptom onset in COVID patients. LIAISON^®^ SARS-CoV-2 S1/S2 IgG had information on the lag time between the onset of symptoms and the tests.They performed serial tests, up to three times, for negative results. We used only the first of the serial measurements in this review. Mount Sinai Laboratory COVID-19 ELISA IgG Antibody Test had information on the timing of sample collection (7 to 14 days after onset of symptoms and retested at 21 days if the initial test was negative). VITROS^®^ Anti-SARS-CoV-2G had information on all patients. But we did not include in our sub-group analysis as their reported time did not fit into our proposed analysis. VITROS^®^Anti-SARS-CoV2T had information on 15 out of 30 subjects. The characteristics of included reports are summarized in table 1. We believe the seven studies provided overall good methodological quality and low risk of bias (Figure 1). See supplemental material for details of the risk of bias assessment (eFigure 2 in the supplement).

**Figure 1.**
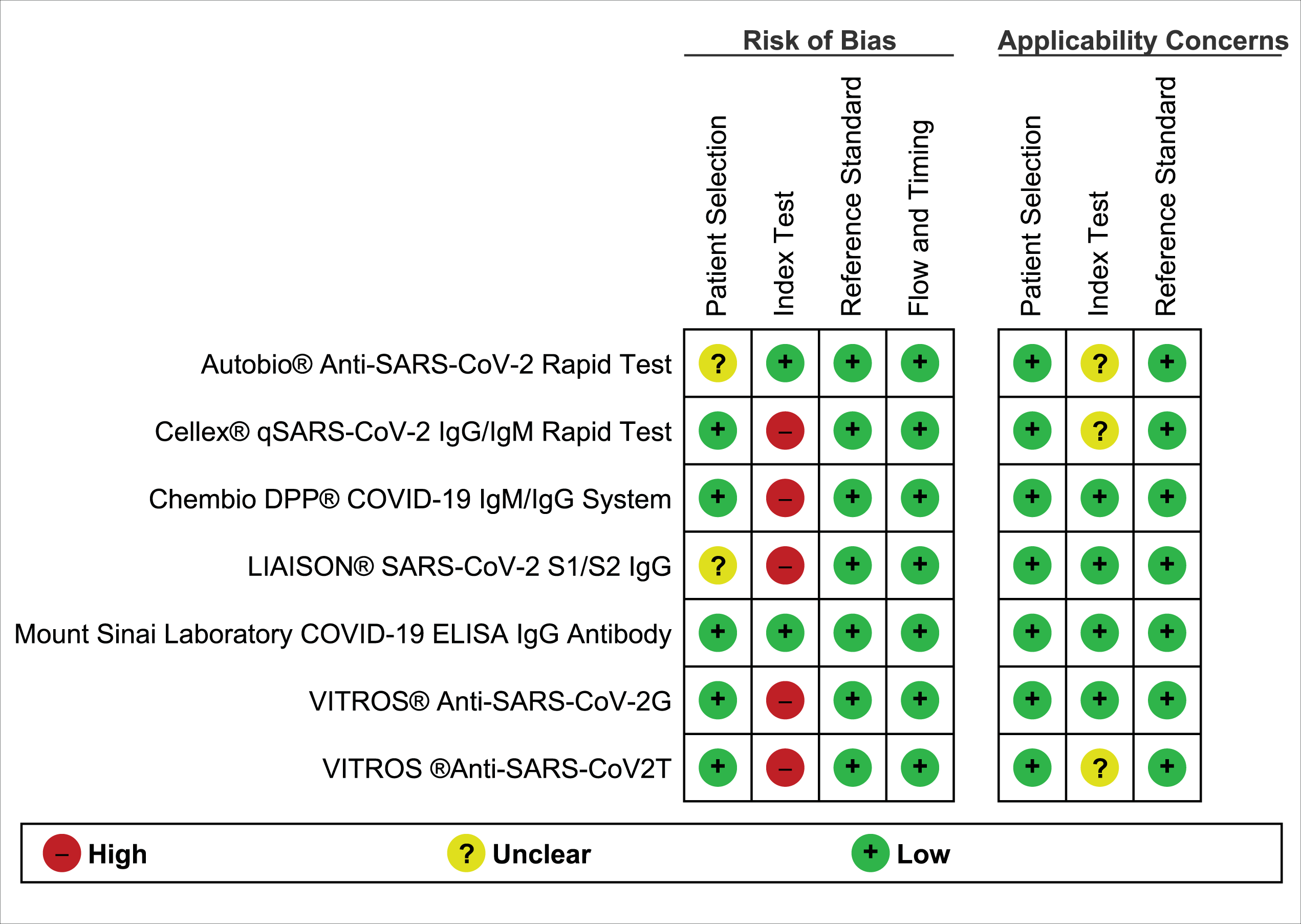
Forest plot: Sensitivity and specificity of serological antibody tests for diagnosing SARS-CoV-2 infection.

### Diagnostic performance of serology tests

The sensitivity of the individual tests ranged from 54% to 94% and specificity ranged from 90% to 100%. We created a coupled forest plot with 95 % confidence interval for individual reports with sensitivity and specificity (Figure 2). The total effect size sensitivity (pooled sensitivity) of all seven studies by the random-effects model was 87% (95% CI, 78% to 93%) (eFigure 3 in the supplement). The Higgins’ I2 was 86%, and the Cochrane Q statistic was 60; p < 0.001, which suggests substantial heterogeneity. The total effect size specificity (pooled specificity) of all seven studies by the random-effects model was 100% (95% CI, 97% to 100%) (eFigure 4 in the supplement). The Higgins’ I2 was 93%, and the Cochrane Q statistic was 26; p< 0.002, which suggests considerable heterogeneity. AUC for the SROC was significant (0.97) (Figure 3). The Spearman’s correlation coefficient between the sensitivity and specificity was negative 0.6, also indicating the presence of an implicit cut-off points. Overall, the diagnostic accuracy of the test was 96% (95% CI, 95% to 97%). The number needed to diagnose was 1.2 (95% CI, 1.1 to 1.2). Around 12 persons need to be tested to return 10 positive tests.

**Figure 2.**
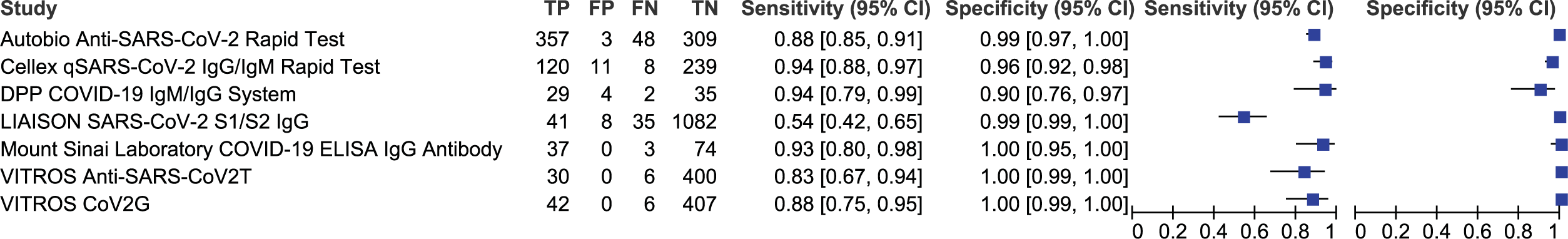
Summary receiver operating characteristic (SROC) plot of serological antibody tests for SARS-CoV-2 infection. Study estimates of sensitivity and specificity are shown with the SROC curve.

**Figure 3.**
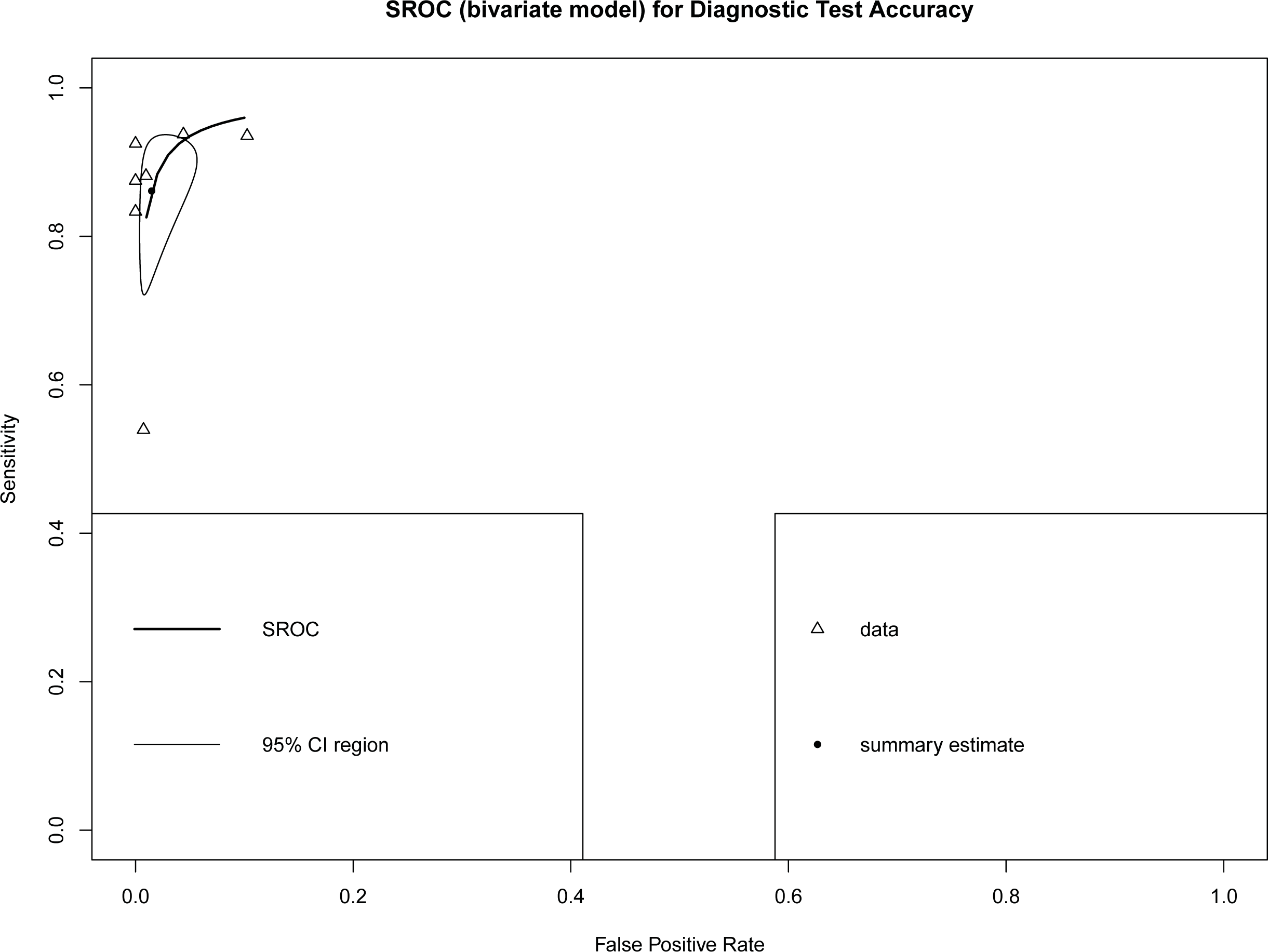
Risk of bias and applicability concerns summary: as judged by the review authors.

### Subgroup analysis

Subgroup analysis detected a notable variation on test sensitivity depending on the timing of blood drawn after the onset of symptoms (lag time). At ≤ 7days (51 samples, 4 reports), sensitivity was 44% (95% CI, 21% to 70%), for 8-14 days (152 samples, 5 reports), sensitivity was 84% (95% CI, 67% to 94%). For blood draws ≥ 15 days after the onset of symptoms (329 samples, 3 reports), sensitivity was 96% (95% CI, 93% to 98%) (Table 2).

**Table 2:**
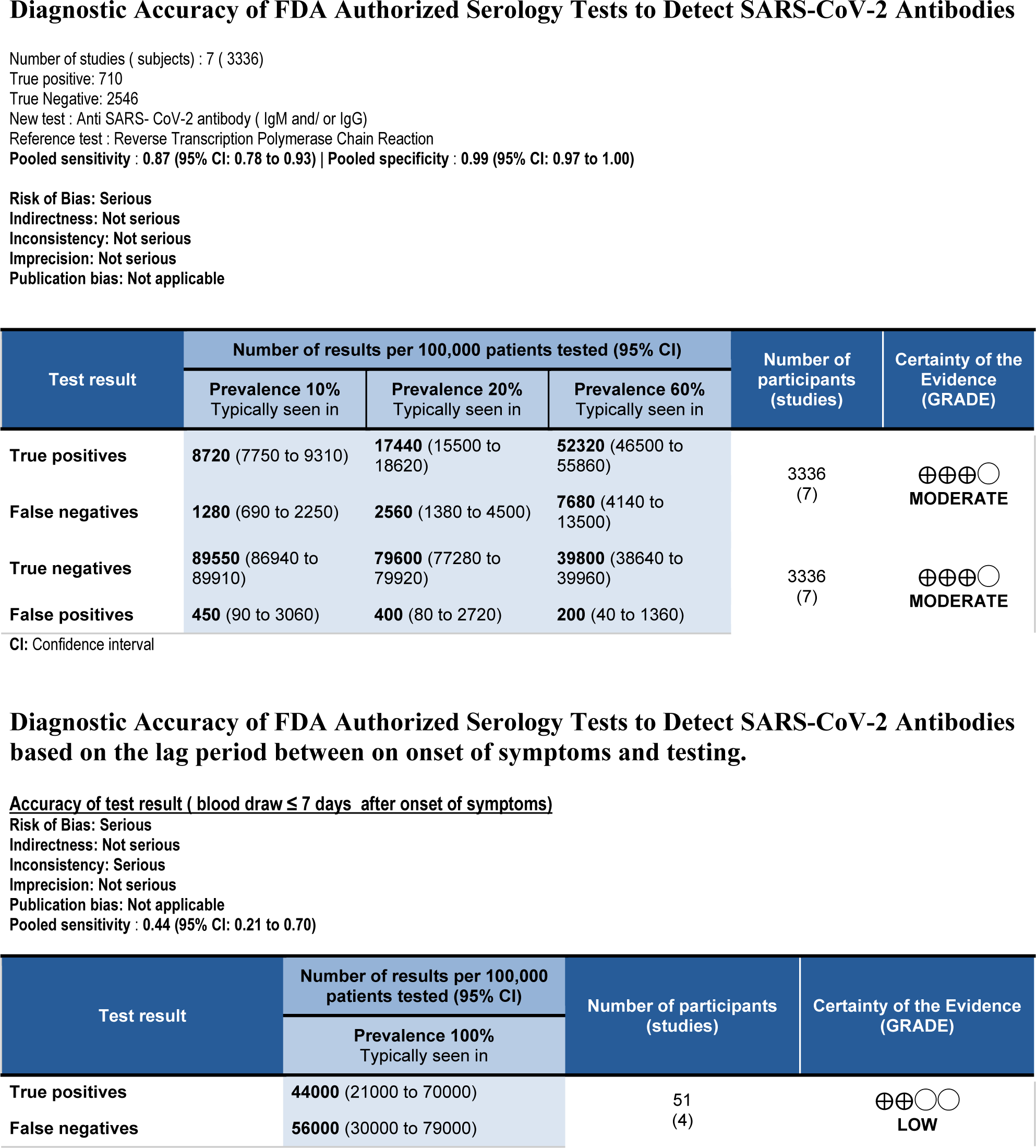
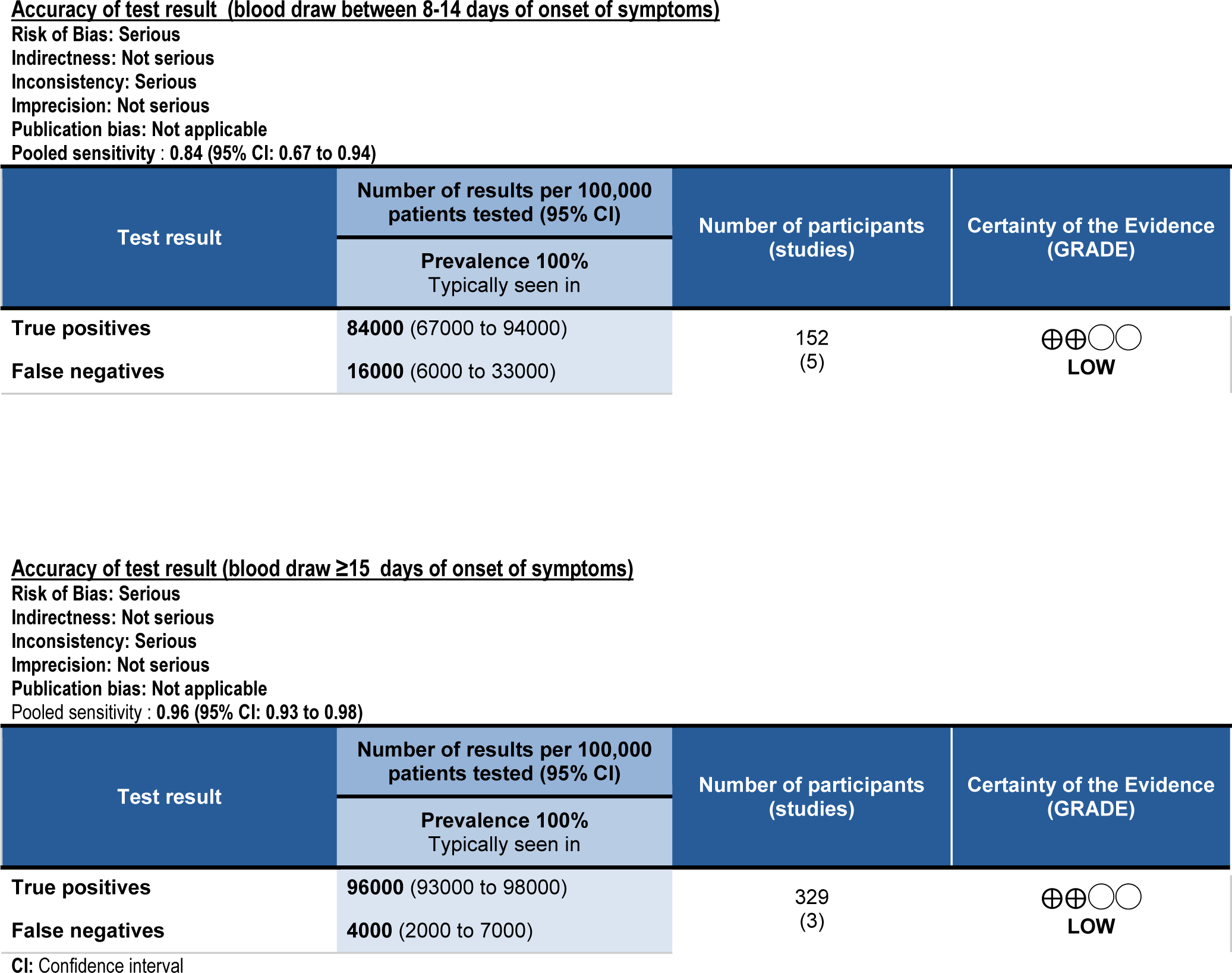
Summary of Findings.

## Discussion

Pooled sensitivity of 87% and specificity of 100 % support that the FDA authorized serology tests are useful in ruling out SARS-CoV-2 infection if the tests are done under similar conditions as conducted in the reports. We noticed variations in testing time from the onset of symptoms as well as serial measurements on some reports. Although heterogeneity of specificity was higher (93%) compared to those of sensitivity (86%), SROC curve with less scattering of test values and moderate Spearman’s correlation coefficient of negative 0.59 validates these studies. The diagnostic accuracy and the number needed to diagnose reflect the strength of the serology tests.

After the onset of infection with SARS-CoV-2, seroconversion occurs variably, ranging from days to weeks. ^27^ Zhao J et al. ^9^ showed empirical evidence for the role of antibody titers with a lag time similar to our study findings. In that study, among 173 patients, the median duration for IgM and IgG antibodies seroconversion was 12 days and 14 days, respectively. Moreover, less than 40% had antibody detected within the first week of illness. In another study of 285 patients, median seroconversion was 13 days for IgG, with 100% positive within 19 days of symptoms. ^19^ IgM antibodies are detectable earlier than IgG antibody (208 plasma sample: IgM median 5 (IQR, 3-6 days) versus IgG median 14(IQR, 10-18 days). ^20^ We found an increase in test sensitivity with an increase in time between the illness and the test [sensitivity 96% for tests done ≥ 15 days from symptom onset (Table 2)]. This high sensitivity finding from only three reports warrants validation from more extensive studies. Negative tests, on appropriate clinical settings, should be interpreted with caution, mostly if the tests are done early in the courses of the disease.

Cross-reactivity of SARS-CoV-2 antibodies with other proteins in the blood is under investigation. Reports of false-positive test results are published. ^28 29^ Cross-reactivity between SARS-CoV-2 antibodies with other known antibodies were tested in all reports. Chembio DPP® COVID-19 IgM/IgG System reported cross-reactivity against human coronavirus HKU1 and human coronavirus 229E. LIAISON® SARS-CoV-2 S1/S2 IgG reported cross-reactivity against the Hepatitis B Virus, Anti-Influenza A, and rheumatoid factors. These cross-reactivity limitations should be considered while interpreting positive tests.

### Limitations

Information on the study population is not available to critically appraise individual reports. We could not verify the spectrum bias. Per protocol, we limited our search to FDA authorized serology tests for detecting antibodies against SARS-CoV-2. None of the serology tests have been independently verified for accuracy by the FDA. Five reports used detection of IgG or IgM or both, and the remaining two used detection of IgG only for the diagnosis of SARS-CoV-2. We accepted the threshold detection of antibodies as defined by individual reports.

Heterogeneity could not be explained by threshold as the serology tests were qualitative. Heterogeneity was substantial. We could not perform meta-regression or sensitivity analysis as limited information was available. Higher heterogeneity in this review can arise from chance, patient selection, and the type of diagnostic test.

We found a significant area under the curve (AUC), which represents the global summary performance of the test. The symmetric shoulder of SROC represents the variability in the studies and the trade-off between sensitivity and specificity.

## Conclusions

WHO and CDC recommend RT-PCR for the diagnosis of SARS-CoV-2 infection. Both CDC and WHO agree that serology tests play a pivotal role in the COVID-19 pandemic considering cost, diagnostic accuracy, and relatively quick results. The overall sensitivity and specificity of the FDA authorized serological tests for detecting SARS-CoV-2 infection are high (certainty of evidence: moderate). There is a wide variation in the test accuracy based on the duration between the onset of symptoms and the tests (certainty of evidence: low).

## Data Availability

This project is based on publicly available data from the FDA website.

## Acknowledgment

Drs. Lisa M. Scheid and Eric W. Reynolds for their review and comments on the draft, and Dr. Audrey Wanger for useful discussions.

